# The NILS study protocol - a retrospective validation study of a preoperative decision-making tool for non-invasive lymph node staging in women with primary breast cancer [ISRCTN14341750]

**DOI:** 10.1101/2022.01.02.22268618

**Authors:** Ida Skarping, Looket Dihge, Pär-Ola Bendahl, Linnea Huss, Julia Ellbrant, Mattias Ohlsson, Lisa Rydén

## Abstract

**Background:** Routine preoperative axillary ultrasonography has proven insufficient for detecting low-burden nodal metastatic deposits. For the majority of newly diagnosed breast cancer patients presenting with clinical T1-T2 N0 disease, the standard axillary staging by sentinel lymph node biopsy is not therapeutic. The pilot non-invasive lymph node staging (NILS) artificial neural network (ANN) model to predict nodal status was published in 2019. The aim of the current study is to assess the performance measures of the model for the prediction of healthy lymph nodes in clinically N0 breast cancer patients at two breast cancer centers in Sweden.

**Methods:** This bicenter, observational, retrospective study has been designed to validate the NILS prediction model for nodal status using preoperatively collected clinicopathological and radiological data. A web-based implementation of the nodal status classifier has been developed and will be used in this study, resulting in an estimated probability of healthy lymph nodes for each study participant. Our primary endpoint is to report on the performance of the NILS prediction model to distinguish between healthy and metastatic lymph nodes (discrimination, N0 vs. N+) and compare the observed and predicted event rates of benign axillary nodal status (calibration).

**Discussion:** Internationally, there are numerous artificial intelligence projects involving non-invasive identification of N0 breast cancer. Here, we present a robust validation study based on external cohorts of our ANN model. Although validation is necessary to show generalizability, it is often overlooked. If the accuracy and discrimination reach a satisfactory level, our prediction tool can be implemented to assist medical professionals and breast cancer patients in shared decision-making on omitting sentinel node biopsy in patients predicted to be node-negative. In future, this may potentially save healthcare resources and reduce costs and adverse side effects. In addition, our study might prompt future studies of nodal metastases of malignancies in other organs, and thus might have implications beyond breast cancer.

**Trial registration:** This study has been prospectively registered in the ISRCTN registry, identification number: 14341750

## Background

Currently, most breast cancer (BC) patients are diagnosed at an early stage of the disease and surgery is the first treatment option for majority of the patients (1). Since early BC is considered curable (2), the number of BC survivors is increasing, and both short- and long-term sequelae from BC treatment are of great importance. It is essential to carefully balance both under- and overtreatment of BC. The benefits of a procedure must outweigh the harms, and we must strive for a high quality of life for BC patients during and after treatment. It is necessary to improve clinical algorithms to choose the most optimal treatment strategy while retaining a high level of safety for the individual patient in front of the treating physician.

The number of nodal metastases is an important prognostic factor for survival in BC, with the most obvious distinction between node-positive and node-negative BC (1). Establishing the correct nodal status for BC patients affects decision-making regarding the use of adjuvant chemotherapy and radiotherapy in early BC (3). However, in the era of personalized treatment, the impact of prognostic information from axillary lymph node status on the medical decision-making process is less important than it was in history, and adjuvant treatment is increasingly tailored towards biological and genetic features of BC (4).

Axillary ultrasound, the most commonly used imaging modality for nodal assessment, is an unreliable staging modality for BC patients with a low nodal metastatic burden; the negative predictive value is not reassuring (5, 6). Thus, for most BC patients presenting with a clinically and radiologically node-negative axilla at diagnosis, in the quest to reliably stage the axilla, the surgical sentinel node procedure is routinely used for staging (7). However, for BC patients aged >70 years with hormone receptor-positive and human epidermal growth factor receptor 2 (HER2)-negative, sentinel lymph node surgery is optional if the patient will receive adjuvant endocrine therapy (7). In the majority (∼80%) of early BC patients undergoing sentinel node surgery, the procedure identifies only healthy lymph nodes with no metastatic tumor deposits (3, 4). For these patients, the invasive surgical procedure is merely diagnostic and not therapeutic.

Although there are many benefits of the minor surgical sentinel node procedure over axillary dissection, sentinel node surgery is associated with short- and long-term side effects. Most importantly, immediate postoperative swelling, paresthesia, arm lymphedema, and self-reported symptoms of arm swelling are documented complications of the sentinel node procedure (8, 9). In addition, the procedure is time-consuming and requires health care financial resources.

We previously developed and published a decision-making tool, the non-invasive lymph node staging (NILS) prediction model, for the prediction of benign lymph nodes in primary BC (10). The NILS prediction model, which is an artificial neural network (ANN) model, was developed using retrospectively collected variables (including patients, N=800). The aim of this study is to retrospectively validate the NILS prediction model in one temporal and one geographical cohort.

## Methods

### Study design

This validation study of the NILS prediction model is a multicenter, retrospective study based on prospectively collected clinicopathological variables included in the patients’ medical charts, mammograms, and pathology reports. In this validation study, we assess the external validity and model validity of the NILS prediction model for healthy lymph nodes and evaluate the discrimination and calibration of the NILS prediction model following the TRIPOD statement (11). A schematic schedule of enrollment, data collection, and monitoring is provided in Figure 1. This study aims to validate the NILS prediction model, a previously published ANN-based decision-making tool, which is based on retrospectively collected data at Lund University Hospital, Sweden. In agreement with the TRIPOD recommendations, the data in this validation study is derived from cohorts different in both time and study sites. Thus, for this validation, patients who underwent surgery for primary BC in Malmö, Sweden in 2020 (N=400, a *temporal* validation cohort) and in Helsingborg, Sweden between 2019 and 2020 (N=200, a *geographical* external validation cohort) will be identified by a national registry of cancer diagnosis/treatment/outcome (INCA). The data retrieved from clinical files will include age at BC diagnosis, medical history of BC, ultrasound assessment of the axilla, and clinical status of the axilla. Variables on tumor size and localization in the breast, including laterality, multifocality, and mode of tumor detection, will be documented from the imaging assessment reports (at first, hand mammography followed by ultrasound assessment in case of an inconclusive mammographic report). In cases where multifocality is detected by either clinical examination or any imaging modality in this study, the tumor will be considered multifocal. Tumor biology data (estrogen receptor status, progesterone receptor status, HER2 status, molecular and histopathological subtype, vascular invasion, and proliferation index Ki-67) will be documented from preoperative core needle biopsy. Number of metastatic axillary lymph nodes will be documented from reports of surgical pathology specimens (sentinel node surgery and axillary dissection).

**Figure 1.**
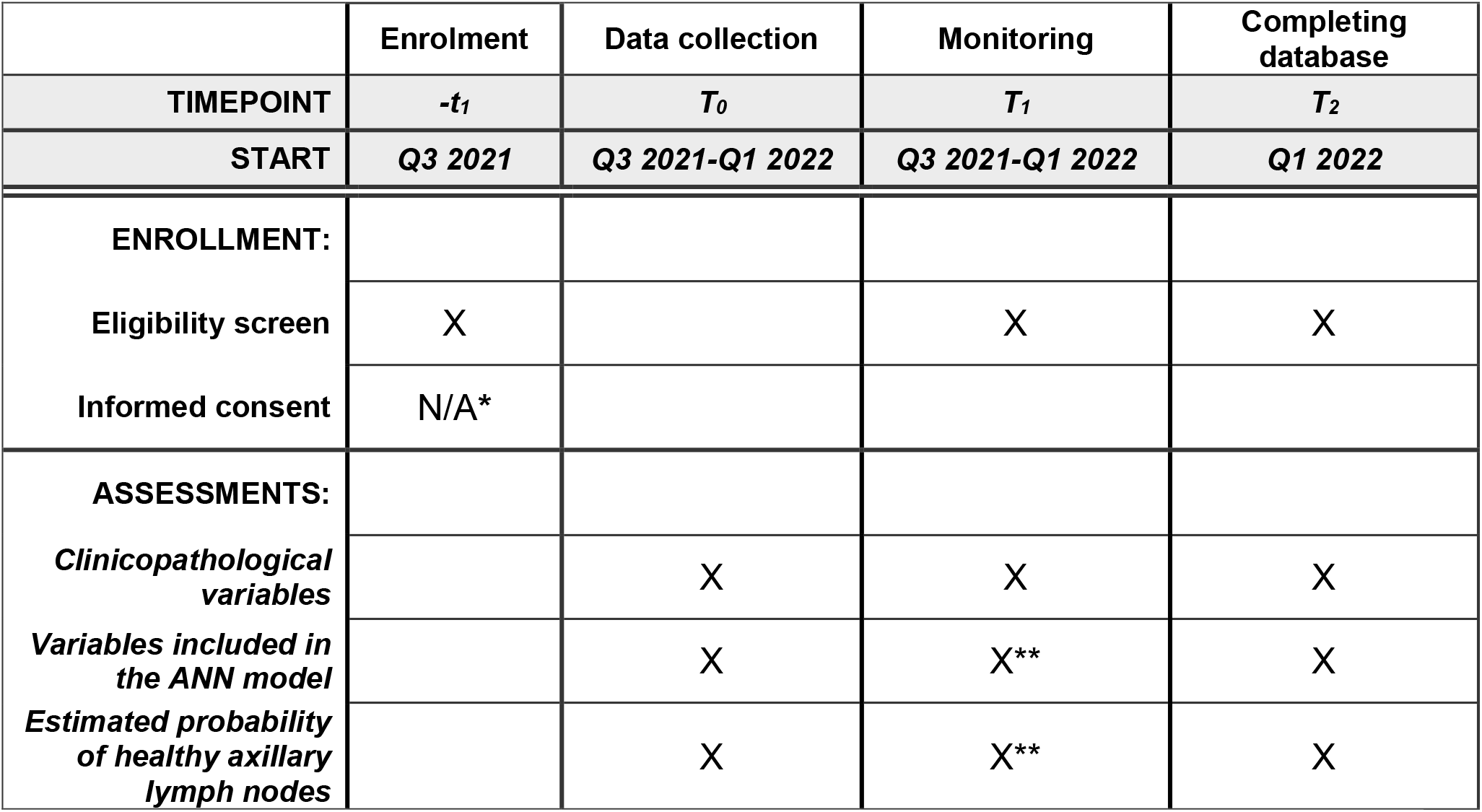
Schedule of enrollment, data collections, and monitoring. *Waived by the ethics committee **A more extensive monitoring schedule for these critical variables (please refer to Supplementary Material 1).

The NILS prediction model for nodal prediction was originally constructed, including ten top ranked risk variables for nodal status (tumor size, vascular invasion, multifocality, estrogen receptor status, histological type, progesterone receptor status, mode of detection, age, tumor localization in the breast, and Ki-67 positivity) (10). The validation study will validate the original model, including the above-mentioned variables. In addition, an important improvement from the early NILS prediction model is that it can also validate the algorithm, including only preoperatively available predictors (i.e., data on biomarkers and histopathological subtypes from core biopsy data and not from breast surgical specimen), and estimated tumor size by imaging rather than by histopathology in the surgically removed tissue.

The decision-making calculator (the NILS prediction model) is derived from an ANN-based model for the prediction of benign lymph nodes at the time of diagnosis in primary BC. The ANN technique can decipher non-linear and difficult-to-predict associations between risk variables and output. In addition, to handle missing values of important variables such as vascular invasion, an additional ANN model that can predict this value using other available variables was developed. A web-based implementation of the NILS protocol, the nodal status classifier, was developed and tested in a pilot cohort of newly diagnosed BC patients. The web interface is being used in this study (Figure 2), resulting in an estimated probability of healthy lymph nodes for each study participant.

**Figure 2.**
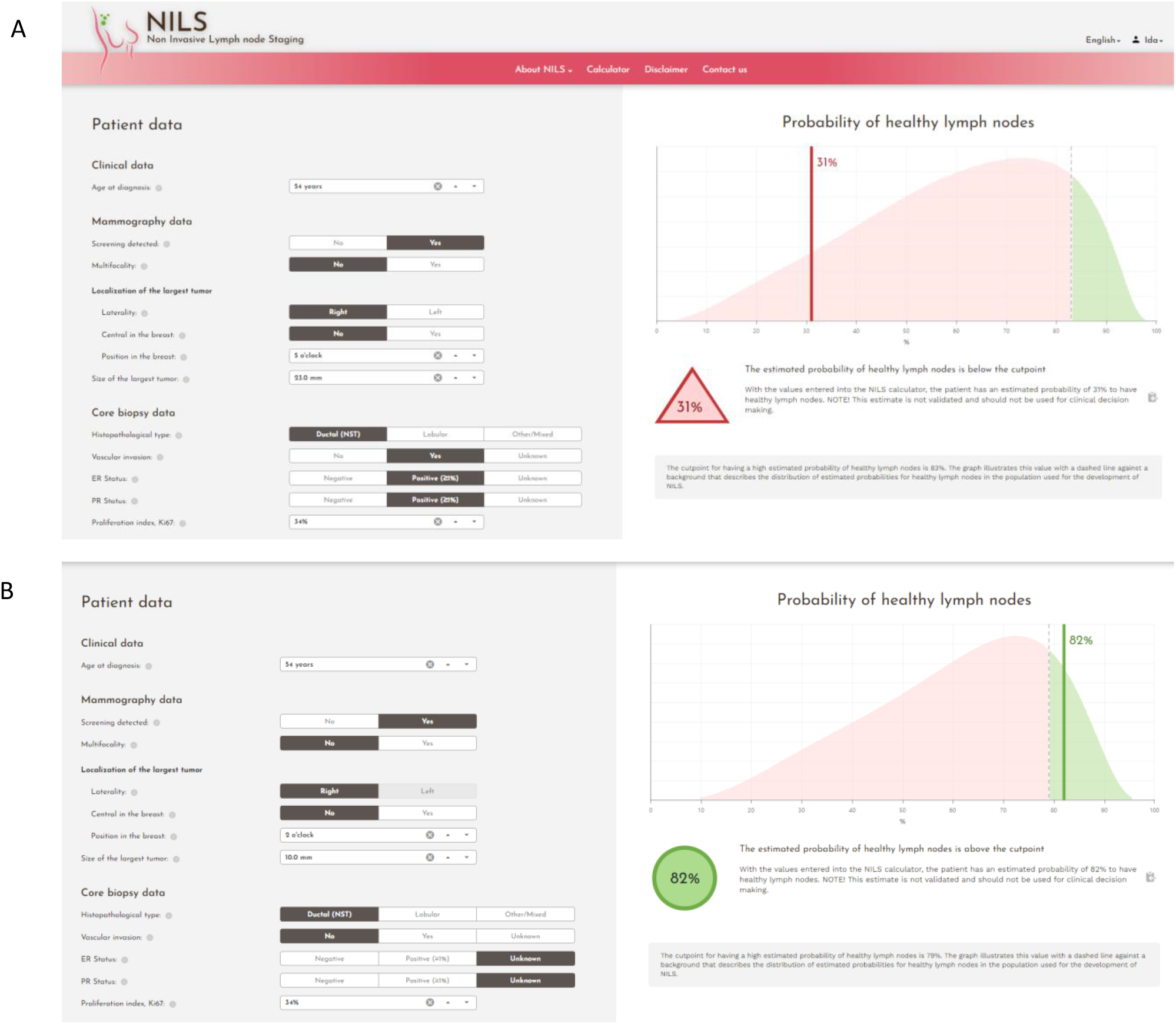
Screen-prints from the web interface of the nodal status classifier. A) Data input resulting in an estimated probability of healthy lymph nodes below the cut point, B) Data input resulting in an estimated probability of healthy lymph nodes above the cut point

This study protocol was written according to the SPIRIT guidelines (12).

### Definition of pN0

In this study, pN0 is defined as sampled axillary lymph nodes with no invasive cell clusters of >0.2 mm at the largest diameter (presence of isolated tumor cells, i.e., tumor cell clusters that are ≤0.2 mm at the largest diameter are considered pN0).

### Ethical approval

All procedures performed in studies involving human participants were in accordance with the ethical standards of the institutional and/or national research committee, and with the 1964 Helsinki declaration and its later amendments or comparable ethical standards. The study was approved by the Regional Ethics Committee in Lund, Sweden (committee reference number: 2021-00174).

The patients received information about the study through advertisement in the local press, and the patients have been allowed to opt out. The ethical committee waived the requirement for informed consent and consent for publication. Upon receiving treatment for BC at the hospitals, the patients gave their consent to register in INCA. The local department for personal data admission at the hospital (KVB Samråd, Region Skåne) granted the researchers access, with digital logging, to the digital medical charts. Access to the web calculator is protected by usernames and passwords, which provides permission to authorized users (physicians, research nurses, and research administrators) from the principal investigator.

### Study population

The included patients had undergone standard diagnostic work-up for the clinical treatment of BC, including a clinical examination, mammogram, ultrasound, and preoperative core biopsy of the tumor. At the time of surgery, the patients underwent standard procedures according to the decision made at the multidisciplinary conference, and the surgical specimen comprising the resected breast tumor and axillary lymph nodes were pathologically examined. No extra diagnostic or interventional procedures were performed within this validation study of a retrospective cohort. No results will be reported to the treating physician or study participants.

### Participating centers

Data is retrieved from two independent centers in southern Sweden: Malmö, a university hospital, and Helsingborg, a regional hospital. There were 0.75 million and 0.3 million people in the catchment area of the breast surgical clinic of Malmö and Helsingborg, respectively.

### Power calculation and statistical plan

We have estimated the sample size to be N=300 study participants, and while each validation cohort will be assessed separately, the total number will be N=600. The area under the curve (AUC) for the original prediction model was 0.74 in the development cohort, where two-thirds of the patients were node-negative (10). Assuming the same proportion of N0 and the same discrimination for future patients, 300 patients are sufficient to obtain 90% probability of reaching an AUC of at least 0.70 in this validation study. This calculation was based on the asymptotic normality of the AUC estimate and simulations that determined the relationship between the sample size and standard error of the AUC estimate. The performance of the ANN-decision tool upon validation will be assessed in terms of discrimination (AUC) and calibration (comparison of the observed and predicted event rates of benign axillary nodal status, Hosmer-Lemeshows test). Separate analyses of patients undergoing breast conservatory surgery and mastectomy will be performed. No interim analyses will be performed. Patients will be screened for eligibility according to specified inclusion and exclusion criteria, and subsequently included if all criteria are fulfilled. Study participants in whom mandatory variables for the NILS protocol are missing, i.e. variables that the ANN model cannot predict itself, will be excluded.

### Inclusion criteria

1. Accepted study participation (e.g. did not opt out)
2. Female
3. Invasive BC
4. Negative preoperative assessment of the axilla, clinically and by ultrasound
5. Scheduled for primary surgery

### Exclusion criteria

1. Did not accept study inclusion (i.e. opted out)
2. Male
3. Children and adolescents (age <18 years)
4. Previous ipsilateral breast/axillary surgery
5. Synchronous distant metastases at diagnosis
6. Previous primary neoadjuvant therapy
7. Preoperative verified axillary lymph node metastases by cytology or histology
8. Positive preoperative assessment of the axilla, clinically and by ultrasound
9. No surgical axillary staging
10. Upfront axillary nodal dissection

### Endpoints

#### Primary endpoint

The primary endpoint is an axillary nodal status (discrimination, N0 vs. N+), determined from preoperative clinicopathological data from mammograms and core needle biopsies compared with the predictive N-status by the algorithm.

#### Secondary endpoint

The secondary endpoint is a false-negative rate of a maximum of 10% for the prediction of N0 vs. N+, compared with the histopathological assessment of excised sentinel lymph node(s).

### Data analysis

For secure and trackable data entry, REDCap (13) with Audit trail is used. A comprehensive data management plan has been developed. Only the principal investigator and responsible researchers will be granted access to the final dataset. Personal information of enrolled patients will be collected, shared, and maintained with a high level of caution and according to local and national regulations to protect confidentiality before, during, and after the trial. All collected data will be stored in an encrypted database (REDCap), which will remain in the custody of the principal investigator.

A risk analysis was conducted, and a detailed plan for data monitoring (conducted by an external researcher) has been established (Supplementary Material 1).

## Discussion

Here, we present a study protocol for the validation of an ANN model for the prediction of healthy lymph nodes in patients with early BC with clinical and ultrasound node-negative disease (ISRCTN14341750).

The sentinel node biopsy technique itself has a well-established threshold for a false-negative rate of 10% (14, 15), and we consider this threshold acceptable in the presented predictive setting of our ANN model. The Swedish Institute of Health Economy has conducted a technical report concluding that our prediction model would be cost-saving if the accuracy is consistent with our previous publication (10).

The optimal management of the axilla in patients with BC is under discussion in the clinical and scientific societies. While a normal standard axillary ultrasound examination is considered insufficient to rule out axillary nodal metastasis (16), improved functional imaging, such as magnetic resonance imaging (MRI) or positron emission tomography/MRI, in the preoperative setting has reached the performance of sentinel node biopsy for excluding axillary lymph node metastases (17-21). Although these imaging modalities have been reported to improve accuracy when determining axillary status, their availability in the preoperative setting is not universal, and the presented data are hitherto based on limited series (Bruckmann *et al*., N=104; Botsikas *et al*., N=80; in a review article by Kuijs *et al*., the largest included study was N=505).

Two randomized controlled trials are investigating the possibility of abstaining from sentinel node biopsy in patients with BC stage T1 undergoing breast-conserving surgery: the SOUND trial (ClinicalTrials.gov Identifier: NCT02167490, estimated data completion Q4 2021) and the INSEMA trial including patients with T1-T2 tumors (ClinicalTrials.gov Identifier: NCT02466737, estimated data completion Q4 2024). The results from these trials will provide qualitative insights into the oncological safety of omitting sentinel node surgery.

Data supporting the impact of pathological staging of the axilla on adjuvant systemic treatment recommendations are limited to luminal A-like tumors, and thus the need to detect axillary lymph node metastasis in early BC is of less importance. In a study by van Roozendaal *et al*., the decision to recommend adjuvant systemic treatment changed in only 1% of the patients due to the pathological lymph node status when using Adjuvant! Online (22), and a report of N=1001 patients included in the INSEMA trial showed that tumor biological parameters alone could guide systemic treatment decisions in 99% of the patients (4).

Many nomograms and ANN models have been developed for predicting nodal metastasis, or the lack thereof, with (23-31) or without imaging (32, 33). The ANN model that this study aims to validate is a promising tool in the clinic because the input variables are routinely available, no extra imaging besides clinical work-up (mammography and axillary ultrasound) is necessary, and the web interface is user friendly.

### Significance

Internationally, there are numerous artificial intelligence projects involving non-invasive identification of N0 BC. The NILS protocol uses easily accessible clinicopathological variables, including mammography data, and in contrast to many other algorithms, it does not require medical imaging data for machine learning analysis. Here, we present a robust validation study of the NILS prediction model based on two external cohorts. If the accuracy and discrimination reach a satisfactory level, our prediction tool (the NILS protocol) can be implemented and would assist medical professionals and BC patients in shared decision-making on omitting sentinel node biopsy in patients predicted to be node-negative. In the future, the use of the NILS protocol can potentially save healthcare resources and reduce costs and unbeneficial side effects. In addition, the NILS prediction model and our presented validation study might prompt future studies of nodal metastases of malignancies in other organs, and thus might have implications beyond BC.

### Current Status

Clinicopathological data are being retrieved from electronic medical charts and pathology reports along with the output from the web application for the ANN. Specially trained research nurses/research administrators with many years of experience in data management are responsible for data entry. Data entry began in Q3 2021 and will be completed in Q1 2022.

## Data Availability

This is not applicable to the study protocol. Following the enrollment of participants, the raw datasets are kept with the corresponding author, and available on reasonable request due to restrictions, such as privacy and ethical restrictions. The data are not publicly available because of these restrictions.

## Declarations

### Ethics approval and consent to participate

All procedures performed in studies involving human participants were in accordance with the ethical standards of the institutional and/or national research committee and with the 1964 Helsinki declaration and its later amendments or comparable ethical standards. The study was approved by the Regional Ethics Committee in Lund, Sweden (committee reference number: 2021-00174).

### Consent for publication

In accordance with the decision of the Ethics Committee an “opt-out” methodology was used and consent for publication was waived by the Ethics Committee.

### Dissemination policy

The study results will be made public through a peer-reviewed publication in an international journal. The full protocol, participant-level dataset, and statistical code will not be publicly available because of privacy and ethical restrictions. Authorship eligibility criteria according to ICMJE recommendations were followed.

### Authors’ contributions

IS and LR drafted the manuscript. LD, POB, LH, JE, and MO substantively revised the manuscript. All the authors approved the submitted version. All authors have agreed to be personally accountable for their own contributions and to ensure that questions related to the accuracy or integrity of any part of the work, even those in which they were not personally involved, are appropriately investigated and resolved, and the resolution documented in the literature.

## Acknowledgements

Many thanks to the research nurses/research administrator Helena Erixon, Eva Wahlström, and Kerstin Reistad in Malmö and Helsingborg for their excellent data management. We also express our gratitude to Forum Söder for the administration of the REDCap project.

## Authors’ information

IS: MD, PhD, Physician at the Department of Clinical Physiology and Nuclear Medicine, Skåne University Hospital, Lund, Lund University, Sweden.

LD: MD, PhD, Consultant reconstructive breast surgeon, Department of Plastic and Reconstructive Surgery, Skåne University Hospital, Lund University, Sweden

POB: PhD, Associate Professor, Statistician at the Department of Clinical Sciences, Division of Oncology, Lund University, Sweden

LH: MD, PhD, Consultant in Surgery, Helsingborg Regional Hospital and Lund University, Sweden JE: MD, PhD, Skåne University Hospital, Malmö, and Lund University, Lund, Sweden

MO: Professor, Department of Astronomy and Theoretical Physics, Division of Computational Biology and Biological Physics, Lund University, Sweden

LR: Professor and senior consultant in Surgery, Lund University and Skåne University Hospital, Sweden

## Table of contents: Manuscript figures and supplementary materials

### Supplementary Material

Supplementary Material 1. Data monitoring

## Abbreviations

BC: Breast Cancer
ANN: Artificial Neural Network
HER2: Human Epidermal growth factor Receptor 2
The NILS protocol: The Non-Invasive Lymph node Staging protocol
MRI: Magnetic Resonance Imaging
AUC: Area Under the Curve

