## Supplementary Material 1 for "The NILS study protocol - a retrospective validation study of a preoperative decision-making tool for non-invasive lymph node staging in women with primary breast cancer [ISRCTN14341750]"

### Supplementary Material 1. Data monitoring – NILS validation study

The first two entered study participants by each data manager will be completely monitored (100%, all data points). Thereafter, data monitoring will be conducted according to the following table. All monitoring will be documented and entered into an Excel file with pre-defined measures of quality.

| **Parameters** | **Extent** |
|---|---|
| Inclusion and exclusion criteria | A cross validation of data reports exported from REDCap (e.g. contradicting data, double data entry, range checks for data values). For all study participants we will establish that complete and correct data is entered. |
| Primary endpoint (pathology N-status *vs.* predicted N-status by the NILS model) | A cross validation of data reports exported from REDCap (e.g. contradicting data, double data entry, range checks for data values). For all study participants we will establish that complete data is entered. |
| All data points | 10% of the study participants will be monitored to 100% (every 10th study participant, starting on a randomly chosen number).<br>If ≤10% errors in non-critical data points → no further monitoring<br>If >10% errors in non-critical data points → increase monitoring to 15% of all study participants for the remaining study participants. |
| Critical points of data (included in the NILS model):<br>• Age at BC diagnosis<br>• Mode of detection<br>• Mulitfocality<br>• Laterality<br>• Position in the breast (central/lateral and clockwise position)<br>• Tumor size by imaging<br>• Histopathological subtype<br>• Vascular invasion<br>• Estrogen receptor status<br>• Progesterone receptor status<br>• Proliferation index (Ki67) | 10% of the study participants will be monitored to 100% (every 10th study participant, starting on a randomly chosen number).<br>If ≤2% errors in critical data points → no further monitoring<br>If >2% errors in critical data points → increase monitoring to 20% of all study participants for the remaining study participants. |
